# The downhill race for a Rainbow jersey. The Epidemiology of Injuries in Downhill Mountain Biking at the 2023 UCI Cycling World Championships using the International Olympic Committee Consensus: A Prospective Cohort Study

**DOI:** 10.1101/2024.02.29.24303534

**Authors:** Thomas Fallon, Debbie Palmer, Xavier Bigard, Niall Elliott, Emma Lunan, Neil Heron

**Affiliations:** Centre for Public Health, Queen’s University Belfast, Northern Ireland; Edinburgh Sports Medicine Research Network & UK Collaborating Centre on Injury and Illness Prevention in Sport (UKCCIIS), Institute for Sport, PE and Health Sciences, University of Edinburgh; Sport Injury Prevention Research Centre, University of Calgary; Union Cycliste Internationale, Aigle, Switzerland; Scottish Institute of Sport, Stirling, FK9 5PH; School of Medicine, Keele University, Staffordshire, England

**Keywords:** injury, surveillance, prospective, cycling, protocol

## Abstract

**Introduction:** Downhill Mountain Biking (DHMTB) is one of the more spectacular sub-disciplines of mountain bike (MTB) cycling. The primary aim of our study was to prospectively document the injury rate, severity, aetiology, location and type during official training and racing by elite DHMTB riders during the 2023 UCI Cycling World Championships.

**Methods:** The participants of this prospective, observational study were elite male and female cyclists competing at the UCI DHMTB World Championships located in the Nevis range in Fort William, Scotland, in 2023. This study followed the injury reporting guidelines established by the International Olympic Committee (IOC), which include the STROBE-SIIS and the cycling-specific extension.

**Results:** Throughout the championships, 10.4% of riders sustained one injury, with 4.3% of riders injuring more than one location per injury event. The overall injury incidence was 3.3 injuries per 100 rides. The incidence rates were higher in the training group (6.4/100rides) than in the race group (2.3/100rides). There was a greater incidence of injury in females in the training 5.7/100 rides and racing 4.4/100rides compared to male riders. Female athletes experienced more severe injuries, with double the estimated time lost to injury. Additionally, female athletes were found to have a significantly greater risk of head injuries and concussions than males.

**Conclusion:** Overall, injuries are more prevalent in training than in competition. Compared with male DHMTB athletes, female DHMTB athletes are more at risk of injury and show a greater incidence of injury within official training and competition as well as more severe injuries.

**Summary Box:** *What is already known:*

- Downhill Mountain Biking (DHMTB) is one of the more spectacular subdisciplines of mountain bike cycling and has been shown to have high injury prevalence.
- There is a lack of methodological homogeneity amongst the prospective injury surveillance studies conducted within DHMTB and across competitive cycling.
- No Study has currently reported injury incidence within elite DHMTB as per the International Olympic Committee (IOC) cycling extension recommendations.

*What this study adds:*

- Within DHMTB injury incident rates were higher in training (6.4/100rides) compared to racing (2.3/100rides).
- Overall Injury incident rate was significantly higher in females (5.1/100rides) compared to males (2.3/100rides).
- Female athletes have a 2.89 higher risk of Injury compared to Male DHMTB athletes.
- Female athletes have significantly higher risk of head/neck (RR 9.5) injuries and concussion (RR 6.34) compared to their male counterparts.

*How this study might affect research, practice, or policy:*

- The IOC Cycling Extension should acknowledge that when reporting injuries per 100 rides, the number of rides completed prior to injury should be collected to improve reporting accuracy.
- Female athletes may benefit from an extra official training ride to ease pressures during course familiarisation and reduce racing injury incidence.
- Female athletes may benefit from neck strengthening and resistance training to reduce the number of head and neck injuries.

## Introduction

The sport of cycling consists of several individual sporting disciplines. The world governing body for cycling, Union Cycliste Internationale (UCI), oversees the cycling disciplines of road cycling, cyclocross, mountain bike (MTB), trail, gravel, BMX freestyle, BMX racing, track, e-sport, para-cycling and indoor. MTB as a discipline has grown exponentially since its founding in 1973 in California to the first world championship in Colorado in 1990 and the Olympics in Atlanta in 1996.[1] Downhill Mountain Biking (DHMTB), one of the more spectacular subdisciplines of MTB cycling, is where riders navigate high-speed, steep, technical descents on rugged trails, aiming for the fastest time to complete the course.

The performance characteristics of DHMTB include rider skill, handgrip endurance, self-confidence, and aerobic capacity.[2] The 2023 UCI Cycling World Championships in Fort William, Scotland, witnessed elite downhill mountain bikers converging to test their skills and mettle on a technically demanding course. Among all levels of competition, the spectre of injuries looms as riders push physical and mental extremes for world champion status, recognised by the awarding of the ‘rainbow jersey’ by the UCI. [3–5]

Across the sport of cycling, injury incidence rates vary by discipline, rider level and study methodology. Numerous studies have examined injury incidences within MTB and have focused on the subdisciplines of MTB. Studies have examined DHMTB,[6–9] cross-country [3, 4, 10–14], enduro [15], MTB stage racing,[16] and MTB park riding.[17] Methodologically, these studies differ from prospective and retrospective studies in terms of design and injury reporting methods. [1, 3, 5, 6, 10, 11, 14–21]

Epidemiological research in DHMTB has shown a unique injury profile characterised by diverse injury types, mechanisms, and anatomical locations.[3, 6] Historically, studies reporting injuries within competitive cycling disciplines lacked methodological guidance and standardised reporting until the publication of the IOC consensus statement extension for competitive cycling in 2021.[22, 23] The IOC consensus statement recommends that DHMTB injuries should be reported per 100 rides, which has not been completed in any DHMTB studies to date.[3, 4, 6] Throughout the sport, cycling lacks high-quality and comparable prospective injury and illness studies across all disciplines. [3, 22] Such research is fundamental in increasing our understanding of risk profiles and possible preventative measures.

The primary aim of our study is therefore to prospectively document the spectrum of injuries incurred by elite DHMTB riders during the 2023 UCI Cycling World Championships in Glasgow, Scotland. This study will extend our knowledge through the comprehensive analysis of injury-related aspects, encompassing characteristics, prevalence, severity, mechanisms, anatomical sites, and the diverse array of factors influencing injury occurrence.

## Methods

The participants of this prospective, observational study were elite male and female cyclists competing at the UCI DHMTB World Championships, located in the Nevis range in Fort William, Scotland, between the 1 and 5 of August 2023. This study followed the injury reporting guidelines established by the International Olympic Committee (IOC), which include the STROBE-SIIS and the cycling-specific extension.[22] Ethical approval for this study was obtained from the Medical Faculty at Queen University Belfast, Northern Ireland (Faculty REC Reference Number MHLS 23_97).

## Implementation

The event medical team received information about the study’s objectives, methodology, and inclusion criteria. All medical personnel were advised to document all injuries that occurred during training and racing from the 1st to the 5th of August at the Championship with the Injury Reporting form (**Supplementary File 1** ). The questionnaire was developed from the consensus on reporting and recording injuries in cycling [22] with injury diagnosis classified by the Orchard Sports Injury Classification System (OSICS) version 10, [24, 25]. The event medical teams were required to complete all injury forms daily and return them to the chief medical officer (CMO) for the event on the same day. The data was screened for duplicates daily, and any incomplete or unclear data was queried (by NH) with the named person completing the form to allow full data capture for the study period. All the cyclists were notified of the study by the event medical staff caring for them and advised that if they objected to their anonymised medical information being shared, they were free to do this, and their medical information would not be shared with the researchers.

## Data collection, inclusion, and exclusion criteria

The questionnaire (**Supplementary File 1** ) was created using a Qualtrics survey-based application by DP with input from NH. All athletes who experienced an injury at the World Championships and sought medical attention were eligible for the study. The inclusion criteria included male or female cyclists in junior and elite competition with no age restriction who competed at the World Championships.

Injury incidence was calculated per 100 rides in training and competition (formula 1) and per 1000 hours in competition (formula 2). As per the UCI rules 4.3.022, “*Riders must complete at least two training runs or they will be disqualified from the race.* [26] *The start commissaire must ensure that this rule is applied.”* Therefore, it was assumed for the calculation of injury incidence rates that each rider completed 4 rides. These are comprised of 2 x official training, a qualification round, and a final round. Total competition exposure time (hrs) was calculated based on the finishing time of each rider in both the qualification and final rounds across all levels obtained from official results.[27]

All data was processed on a Macintosh computer using Microsoft Office and SPSS (V.28). The methods applied included frequencies (%), crosstabs and descriptive statistics. The relative risk (RR) was calculated using formula 3 and is presented with 95% confidence intervals (CIs). All the cyclists were analysed together, and the different cycling demographics (gender, level, injury severity, and training vs. racing) were analysed separately to allow for the comparison of injury data between the disciplines. All the statistical tests were two-sided, and results with p<0.05 were considered to be statistically significant.

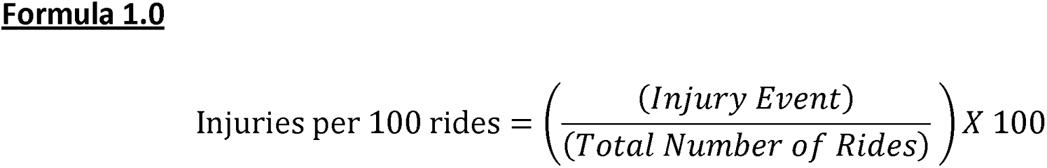

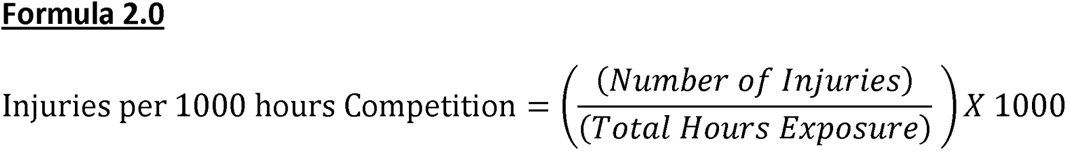

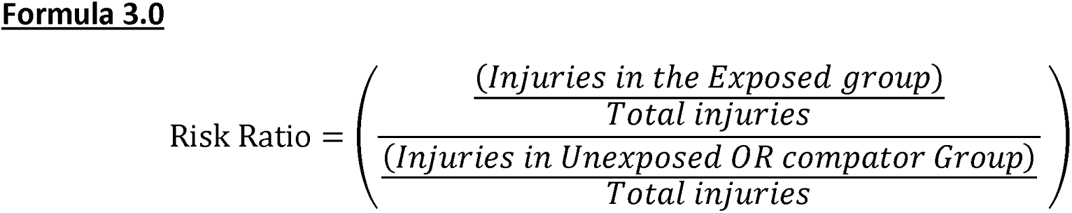

## Results

A total of 230 riders (152 male; 78 female) competed in the 2023 elite DHMTB world championships. A breakdown of the rider demographics is presented in Table 1.

**Table 1.**
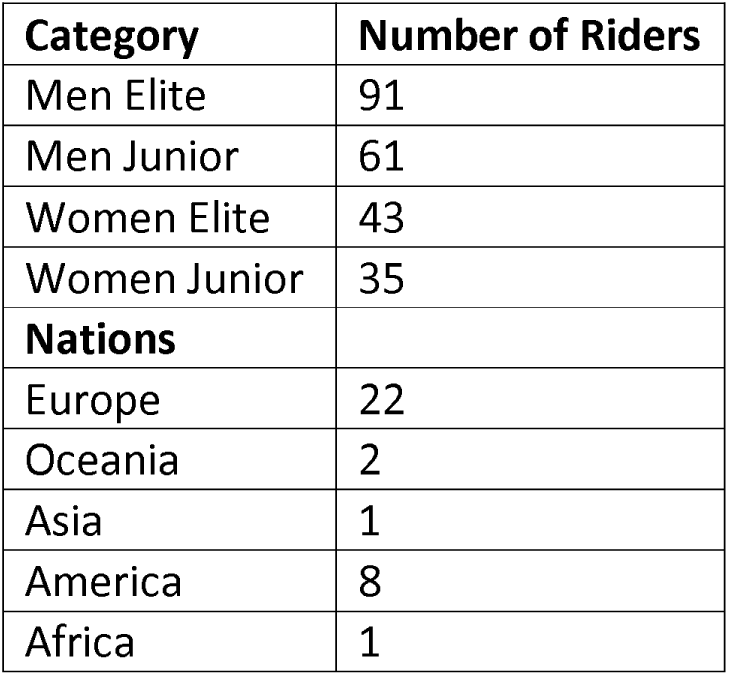
Event Rider Demographics.

| **Category** | **Number of Riders** |
| --- | --- |
| Men Elite | 91 |
| Men Junior | 61 |
| Women Elite | 43 |
| Women Junior | 35 |
| **Nations** |  |
| Europe | 22 |
| Oceania | 2 |
| Asia | 1 |
| America | 8 |
| Africa | 1 |

Overall, 10.4% (n=34) of all riders sustained at least one injury during the 2023 UCI DHMTB World Championships. Of these, 4.3% of riders injured more than one body location on one ride, with 70% of these injuries occurring in female riders. During the world championships, 59% of the injuries occurred in female athletes, and 65% of all injuries occurred in non-professional riders. However, there was no significant difference between the number of years of racing and injury locations (p=0.128).

The overall event injury incidence rate was 3.3 injuries per 100 rides. During training, there were 4.3 injuries per 100 rides, and in competition, there were 2.2 injuries per 100 rides (**Table 3**). Expressed relative to the number of competition hours, the injury incidence was 7.6/1000 hrs (**Table 3**). The average estimated time lost due to injury was 5.5 (±1.6) days for male riders, for a maximum of 14 days. Female athletes lost more than double the estimated time due to injury, with an average of 12.6 (±14) days and a maximum of 42 days. There was no significant difference between the level of athlete (p=0.239) or sex (p=0.445) and the time lost to injury (**Table 2,4**).

**Table 2:** Percentage of injuries (and total number in brackets) reported within training and competition, rider level and sex.

| **Injuries** | **Male (%)** | **Female (%)** |
| --- | --- | --- |
| In Competition | 21 (3) | 55 (11) |
| In Official Training | 79 (11) | 45 (9) |
| **Level of cyclist** |  |  |
| Non-Professional | 57 (8) | 70 (14) |
| Professional | 43 (6) | 30 (6) |

**Table 3:** Injury incidence rates and severity (days)

|  | **Inj/100 Rides (95%CI)** | **Injury Severity (95%CI)** | **Inj/1000 Hours (95%CI)** |
| --- | --- | --- | --- |
| Training Overall | 4.3 (4.0 – 4.6) | 6.3 (3.13-9.47) | - |
| Racing Overall | 2.2 (2.1 – 2.3) | 14.5 (4.8- 24.2) | 7.6 (7.3- 7.9) |
| Male Training | 3.6 (3.3 – 3.9) | - | - |
| Male Racing | 0.94 (0.12-1.2) | - | 1.13 (1.1- 1.2) |
| Female Training | 5.8 (5.0 -6.6) | - | - |
| Female Racing | 4.5 (3.9- 4.9) | - | 14.2 (13.2- 15.2) |
| Female overall | 5.1 (4.6 - 5.6) | 12.60 (5.66-19.54) | - |
| Male Overall | 2.3 (2.2 - 2.4) | 5.50 (1.89 – 9.11) | - |
| Overall | 3.3 (3.1- 3.5) | 9.68 (5.34-14.01) | - |

**Table 4:**
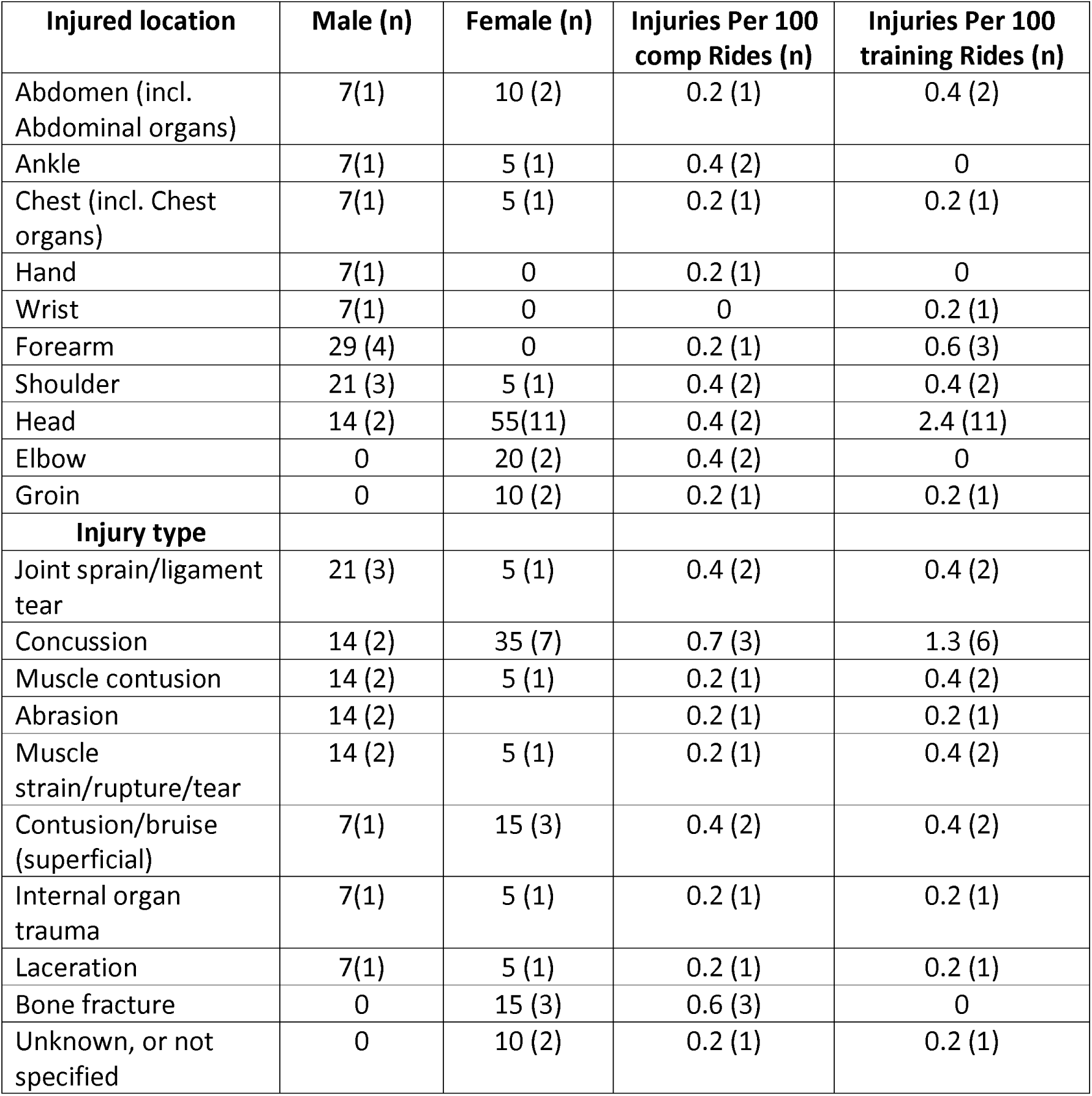
Percentage of Injuries by location, injury type and incident rate (with the number in parentheses)

| Injured location | Male (n) | Female (n) | Injuries Per 100 comp Rides (n) | Injuries Per 100 training Rides (n) |
| --- | --- | --- | --- | --- |
| Abdomen (incl. Abdominal organs) | 7(1) | 10 (2) | 0.2 (1) | 0.4 (2) |
| Ankle | 7(1) | 5 (1) | 0.4 (2) | 0 |
| Chest (incl. Chest organs) | 7(1) | 5 (1) | 0.2 (1) | 0.2 (1) |
| Hand | 7(1) | 0 | 0.2 (1) | 0 |
| Wrist | 7(1) | 0 | 0 | 0.2 (1) |
| Forearm | 29 (4) | 0 | 0.2 (1) | 0.6 (3) |
| Shoulder | 21 (3) | 5 (1) | 0.4 (2) | 0.4 (2) |
| Head | 14 (2) | 55(11) | 0.4 (2) | 2.4 (11) |
| Elbow | 0 | 20 (2) | 0.4 (2) | 0 |
| Groin | 0 | 10 (2) | 0.2 (1) | 0.2 (1) |
| **Injury type** |  |  |  |  |
| Joint sprain/ligament tear | 21 (3) | 5 (1) | 0.4 (2) | 0.4 (2) |
| Concussion | 14 (2) | 35 (7) | 0.7 (3) | 1.3 (6) |
| Muscle contusion | 14 (2) | 5 (1) | 0.2 (1) | 0.4 (2) |
| Abrasion | 14 (2) |  | 0.2 (1) | 0.2 (1) |
| Muscle strain/rupture/tear | 14 (2) | 5 (1) | 0.2 (1) | 0.4 (2) |
| Contusion/bruise (superficial) | 7(1) | 15 (3) | 0.4 (2) | 0.4 (2) |
| Internal organ trauma | 7(1) | 5 (1) | 0.2 (1) | 0.2 (1) |
| Laceration | 7(1) | 5 (1) | 0.2 (1) | 0.2 (1) |
| Bone fracture | 0 | 15 (3) | 0.6 (3) | 0 |
| Unknown, or not specified | 0 | 10 (2) | 0.2 (1) | 0.2 (1) |

Compared with male athletes, female athletes were at significantly greater risk of overall injury (RR 2.21, 95% CI 1.5-5.4). Female athletes were found to have a significantly greater number of head injuries (RR of 9.5, 95% CI 2.15-41.9 p=0.002) and incidence of concussion (RR of 6.34, 95% CI 1.34-29.84, P=0.01) when compared to males. Also, those diagnosed with concussion had a greater prevalence of headache/neck pain (**Figure 1** ). All bone fractures were confirmed in non-professional female athletes. Female athletes presented a greater number and prevalence of observable concussive signs (**Figure 1**).

Among male riders, 50% (n=7) of injuries led to no time off. Among female athletes, 40% (n=8) of the injuries that required medical attention led to no time off. The mechanism of injury and removal from the racing status are presented in **Table 5**.

**Table 5:** Percentage of injuries by mode of onset, injury mechanism, and injury burden (number of injuries)

| Cycle specific injury mechanism | Male (n) | Female (n) |
| --- | --- | --- |
| Collided with an inanimate object related to competition | 7.1 (1) | 5 (1) |
| Collided with the ground only | 92.8 (13) | 95 (19) |
| **Injury mechanism** |  |  |
| Direct contact with an object (e.g. Ball, wall, ground, i.e., Slipped and fell) | 100 (14) | 100 (20) |
| **Unable to finish and needed to be removed from field of** |  |  |

| **play** |  |  |
| --- | --- | --- |
| No | 64 (9) | 45 (9) |
| Yes, removed from play | 35 (5) | 55 (11) |

## Discussion

This was the first study within DHMTB to apply the IOC consensus statement extension for the reporting of injury and illness in competitive cycling. [22] The study aims were to describe and compare the injury rate, severity, aetiology, location, and type of injury during official training and competition at the 2023 DHMTB UCI World Championships at the Nevis range in Fort William, Scotland. The main findings were as follows:

1) Throughout the championship, 10.4% of riders sustained at least one injury, with 4.3% of riders injuring more than one body location per incident;
2) The overall injury incidence rate was 3.3 injuries/100 rides;
3) Injury incidence rates were greater during training (4.3/100ride) than during competition (2.2/100ride);
4) The injury incidence rate was significantly greater in females.
5) Females had a 6.3-fold greater risk of concussion injury and a 9.5-fold greater risk of head/neck injuries than males did.

### Injury Rates

To date, few prospective studies have examined injuries within DHMTB.[3, 4, 6] Our study revealed a relatively low incidence of injury across the World Championship DHMTB compared to that reported in previous DHMTB studies [3, 4], those reported in the DHMTB World Cup series (80%) [6], cross-country MTB events[16] (71%) and MTB at the London (16%) and Rio (24%) Olympic games.[11, 12] However, the 10.4% overall incidence of injury observed was higher than the 8.4% observed in enduro cycling and 7% in MTB at the 2021 Tokyo Olympics [10, 15]. Consideration must be given to the similarities and differences in risks and demands between MTB disciplines when comparing injury proportions.

One of the unique strengths but also challenges of this study is the expression of injury incidence rates. The IOC consensus cycling extension recommends that injury incidence rates be expressed per 100 rides in DHMTB, in addition to reporting incidences per 1000 hrs of exposure within competition.[1, 3, 4, 6, 16] In training, injury incidence rates of 1.08/1000 hrs within DHMTB world cups [6] and 3.6/1000 hrs in enduro cycling [15] have been observed. Athletes included in these studies would have encountered similar timed downhill sections as athletes in this study. Our findings of injury incidents in racing are lower than those noted in previous DHMTB studies of 16.8/1000 hrs [3] and 43/1000 hrs [4] and within enduro of 38.3/1000 hrs [15]. Limited comparisons can be made between injury incidence in training or racing and previous DHMTB studies because they were not differentiated [3] or included [4] within previous surveillance captures. However, the overall injury incidence trend differs from that in field-based sports, such as amateurs [28], premiership [29] rugby and football [30], where injury incidences are greater in competitions than in training.

Injury incidence rates can be influenced by many factors. Like enduro cycling, DHMTB requires technical skill, speed, concentration, reaction, aerobic capacity, and strength.[2] Thus, to win at the elite level, athletes are pushed to the limit physically and mentally. The level of performance (recreational vs elite, regional vs national, national vs international), duration of the course, and technical nature of the course influence the challenge of different skill profiles and influence injury incidence.[3, 4, 6] Furthermore, the methodology of the study will influence incident rates, i.e., retrospective vs prospective reporting,[3, 6, 15, 31] or self-reported vs physician-diagnosed [1, 3, 15, 17], which could increase the risk of bias.[32, 33] Few studies have followed the IOC consensus [22, 23] when reporting injuries and illness in MTB [3, 11–13, 15, 19], with this being the first study to do so within elite DHMTB. [21] Our study supports the need for further surveillance research that follows the IOC consensus recommendations within cycling.[22]

### Influence of Sex on Injuries

This is the first study to present differences in injuries between sexes among elite DHMTB athletes. Overall, female athletes exhibited significantly greater incidence rates than males did, with an RR of 2.89 (95% CI 1.5-5.4). Despite the higher incidence of injury observed in competition, female athletes’ injury incidence rates (4.5/100 rides (14.2/1000 hrs) and injury RR (2.0, 95% CI 0.64-6.23) did not significantly differ from those of males (0.94/100rides (1.3/1000 hrs)). These findings are similar to those of previous MTB studies [15] and injury incidence rates in team-based sports.[34] Female athlete participation has grown in the past decade, and studies within DHMTB date back to 1996[4]; more recent studies in DHMTB did not include sex-specific analysis,[3, 6] potentially limited by the competitive sample of female athletes. A limitation when presenting DHMTB injury incidence per 100/rides is that both male and female riders compete on the same course. Male riders complete the course quicker than their female counterparts do; therefore, the duration of risk exposure for females is greater than that for males, which is not represented per 100 rides. When comparing the injuries in racing per 1000 hrs, our overall injury incidence rates (7.6/1000 hrs) are lower than those seen in previous studies in the DHMTB[4, 35] and enduro[15]; however, they are similar to those seen within elite competition road cycling studies.[36, 37] The greater injury trends among female athletes observed in this study are similar to those observed in prospective cycling studies of pro-enduro athletes [15]. However, these sports differ from team-based sports in that there is no difference between male and female injury incidence.[38] Our findings raise the question of whether additional work needs to be done to protect female DHMTB cyclists. Examples of injury prevention could include rule changes, for example, adding an extra official practice ride in training or introducing injury prevention programs, such as similar programs to the FIFA 11+ or Gaelic Athletic Association 15, particularly in terms of neck strengthening and concussion prevention.[39–41]

### Injury regions and injury types

Our findings highlight the spectrum and prevalence of injuries among elite male and female DHMTB cyclists. Among male DHMTB riders, joint sprains and ligament tears were the most common, with 21% of injured males reporting such injuries. These injury regions are like previous cycling studies in the DHMTB, enduro MTB and cross-country MTB showing that joint and ligament injuries have a high prevalence within males.[3, 4, 10–12, 15, 35] The high prevalence of forearm (29%) and shoulder (21%) injuries observed in male DHMTB may be explained by protective fall behaviour displayed by male athletes. Sport-specific steps simulating injury prevention behaviour in training may aid coaches and medical professionals in educating athletes on fall techniques to reduce the injury severity from falls in the DHMTB.[42]

Bone health in cyclists has been a key topic of discussion throughout the past decade.[43] However, inconsistent data has shown a relatively high prevalence of low bone mineral density among cyclists, with an increased prevalence seen in elite road cyclists.[44–46] Arguably, elite road cyclists are more at risk of low bone mineral density due to a greater risk of relative energy deficiency (REDs) than DHMTB cyclists, which is more explosive.[47] Early-career female cyclists had lower bone mineral density compared to their male counterparts, which may influence fracture risk with acute injury.[48] All fractures observed in this study occurred in non-professional-level female athletes. Many factors, such as physical, and technical capabilities, may explain the greater fracture injury risk noted among amateur DHMTB cyclists than among their professional counterparts. Additionally, the small sample size contained within this study will impact the strength of the associations observed.

Concussion accounted for 24.5% of all injuries, with a 35% prevalence among female DHMTB athletes. The overall incidence of concussion observed in the DHMTB cohort was similar to that observed in previous DHMTB studies (25% [35] and 23.6%, respectively) [19]. However, the reported incidences of concussion vary significantly within MTB cycling, with an incidence as low as 5% being reported; this incidence is arguably linked to diagnostic challenges and variability in course styles and competition demands across MTB disciplines.[5, 35, 49] Some authors have reported that female athletes are more susceptible to concussion and have more prolonged symptoms after concussion.[50] Among female cyclists, 3.7/1000 hours had a concussion incidence rate within the race, similar to the overall 3.9/1000 hours seen in enduro racing.[15] Our injury incidence rates among female cyclists are similar to those seen in female footballers (3.5/1000 hrs)[51] and slightly greater than those seen in female rugby 15 players (2.8/1000 hrs).[52] However, our findings are much lower than those observed for the rugby 7 s (8.9/1000 hrs) and rugby league (10.3/1000 hrs) plants.[52]

The most reported concussion symptoms were headache and neck pain, with visual problems/amnesia and disorientation commonly reported among both male and female riders. These symptoms are included in the recently published SCAT-6 symptom checklist [53] and the UCI concussion recognition pocket tools [54]. There have been recent calls for action around the diagnosis of concussion within DHMTB.[49] Our findings highlight a lower incidence of concussion injury (n=3) in racing compared to training (n=6), arguably linked with the diagnostic challenges of making this diagnosis.[49] Our findings raise the question of whether there is a link between neck strength and concussion in individual sports.[55] Compared with male DHMTB athletes, female DHMTB athletes were found to have a significantly greater RR of neck injury. There is a weak relationship between neck strength and concussions in team-based sports, and research has shown that female athletes have 47% less neck strength than males.[50, 55, 56] Like male DHMTB cyclists, female DHMTB cyclists use similar-weight helmets, and their reported risk of crashing is almost double the amount of neck pain/headache. During crashing, the differences in neck strength between female athletes and helmet weight may add to the peak linear acceleration and rotational sheer, leading to whiplash injury (neck pain) and potentially a coup contra coup mechanism leading to concussive symptoms.[57] Although no studies have been completed to date in cycling, sports such as ice hockey have linked helmet geometry and injury mechanisms to play a role in concussion.[58] This requires further exploration within DHMTB.

### Implications for Injury Prevention within Cycling and DHMTB?

The epidemiological insights presented within this study will provide a foundation for the discussion and subsequent development of evidence-based injury prevention and management strategies within the DHMTB. As this was the first study to apply the IOC cycling extension to injury, it has raised points of improvement around the consensus application in cycling. When reporting injuries per 100 rides or 100/rounds, the round in which the injury occurred should be included to improve the accuracy of the data. This point should also apply to some events in BMX and track cycling sprint events. Additionally, within the DHMTB, our study showed that the incidence of injury during practice was greater than that during racing in both males and females. As per UCI rule 4.3.022, there is a prerequisite requirement to officially ride 2 practice rounds.[26] For male riders, this is satisfactory given the low incidence of injury observed within racing. However, for female athletes, should there be a protected time window for an additional practice round or mandatory review of the course video analysis, which may reduce the number of crashes/injuries seen in racing? With the high prevalence of neck/head injuries, including concussions, noted in female DHMTB athletes, the inclusion of neck strengthening exercises may have a positive impact on reducing soft tissue injury risk as well as reducing concussion risk. Last, the high incidence (15%) and RR (13.5) of bone fractures noted among female athletes raise the importance of encouraging resistance-based exercise and screening for risk factors for low BMD in cyclists. These risk factors include low body mass index, fracture incidence, smoking, lack of bone-specific physical activity, and low energy availability. [48]

### Limitations and Future Considerations

Given that this study was descriptive and included small sample size, the associations noted cannot be assumed to be causative factors for injury. Second, as this was a “within competition surveillance study”, the methodology of this study was biased toward acute injuries; thus, the representation of illnesses and overuse injuries will likely be underreported. Last, in contrast to racing, determining rider training exposure (hours) directly was not feasible. Therefore, a composite indirect measure using the number of rides in line with the rules was used.

## Conclusion

To our knowledge, this is the first study to prospectively examine injuries within the UCI DHMTB World Championships and report these injuries in line with the cycling extension of the IOC consensus statement. This study provides insights into the injury trends that athletes are exposed to at world championships and can be used to inform injury prevention programs and basis to recommend rule changes in the future. Compared with male DHMTB athletes, female DHMTB athletes are significantly more at risk of injury than their male counterparts are at risk of injury, and they show a greater incidence of injury within official training and competition, particularly in terms of greater risk of head injury and concussions. This study further highlights that injury surveillance can be performed with little burden on event organisers and medical staff and calls for the UCI to endorse similar projects at major competitions. This approach will facilitate the development of our knowledge of the prevalence of injuries within various cycling disciplines.

**X (Twitter)**: @TFallon_Physio, @UKCCIIS, @neilsportsdoc,

## Contributors

The authors would like to thank the Nevis Range medical team for their help with the study in completing the injury reporting forms. Additionally, we thank the UCI and Glasgow 2023 World Championship medical team for endorsing the study.

## Funding

TF is funded by the Department for Education (DfE). No other funding was received for this study.

### Competing interests

None declared.

**Supplemental material** This content has been supplied by the author(s):

**Figure 1.** Concussion Symptom prevalence between Female and Male athletes.

**Sup File 1:** Appendix 1: Survey Log.

**Sup File 2:** Appendix 2: STROBE Statement—Checklist of items that should be included in reports of *cohort studies*.

***Sup File 3:*** Appendix 3: CHAMP: CHecklist for statistical Assessment of Medical Papers

## EDI Statement

Our authorship team included two women and four men, with a mix of senior and early-career research experience. Additionally, the team cover a variety of disciplines (musculoskeletal physiotherapy, General Practice, sport and exercise medicine, and epidemiology) with specialist knowledge, clinical experience, and interest in cycling medicine.

## Data Availability

All data produced in the present study are available upon reasonable request to the authors

## Appendix 1: Survey Log

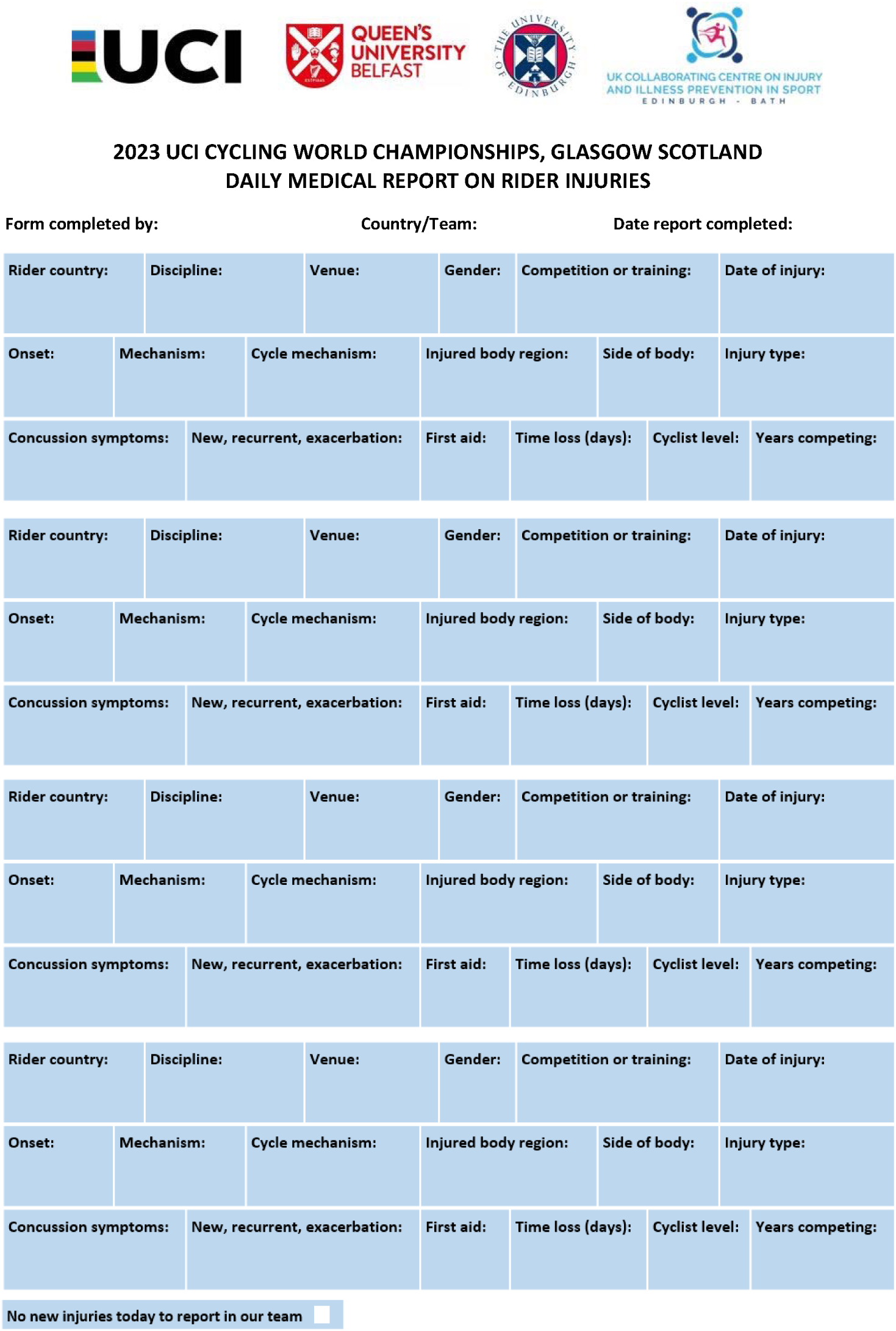
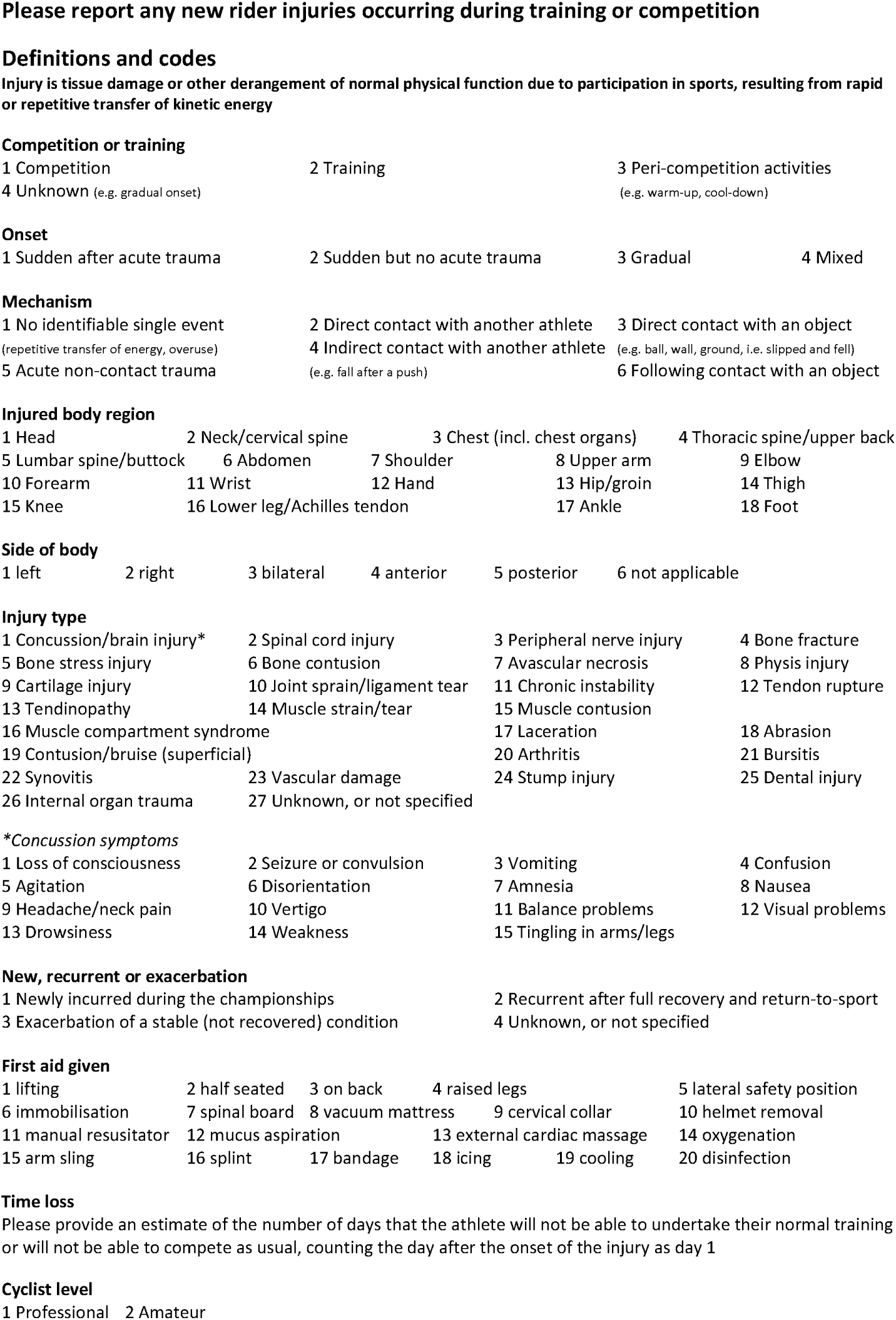

## Appendix 2: STROBE Statement—Checklist of items that should be included in reports of *cohort studies*

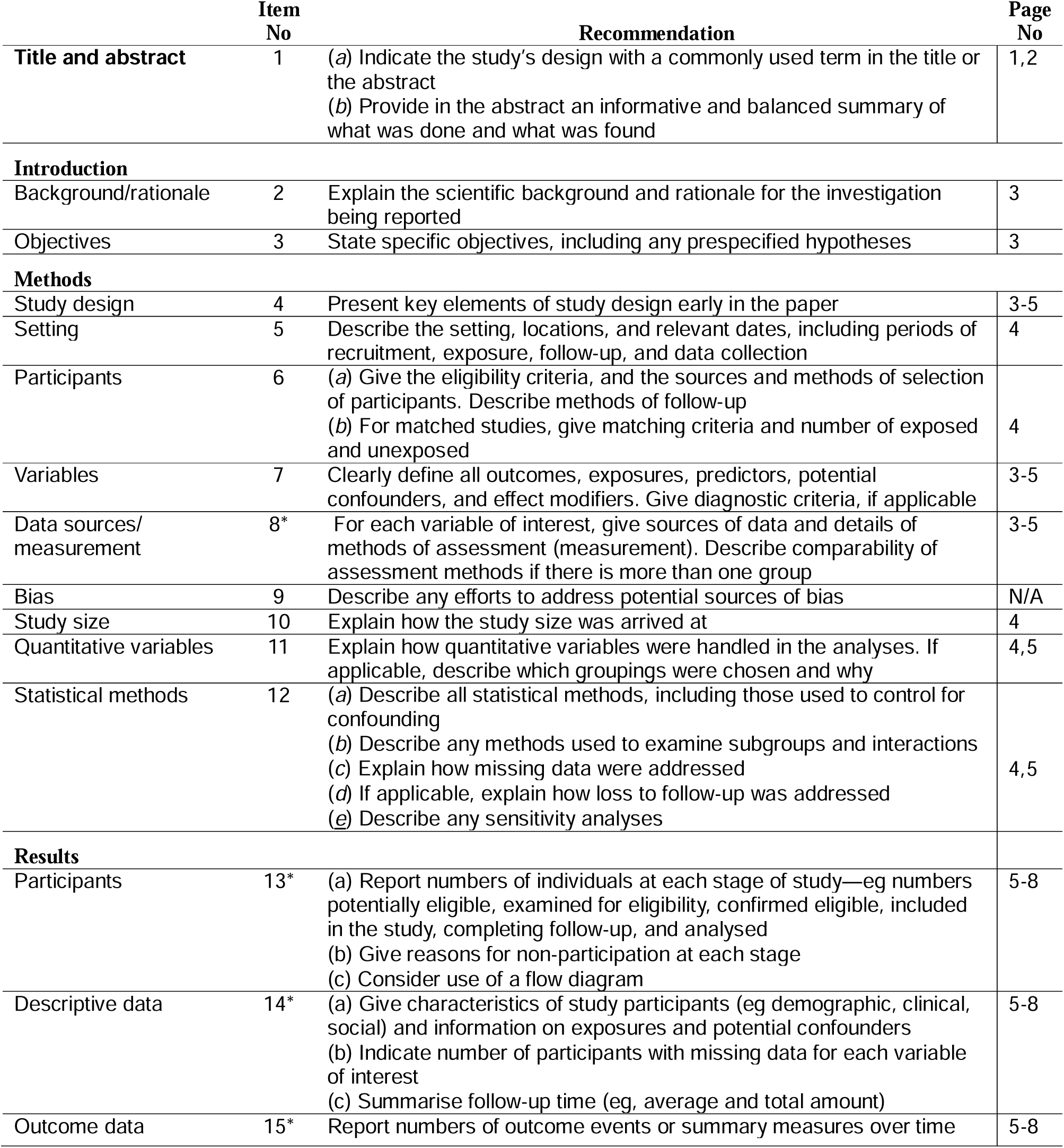

## Appendix 3 CHAMP: CHecklist for statistical Assessment of Medical Papers

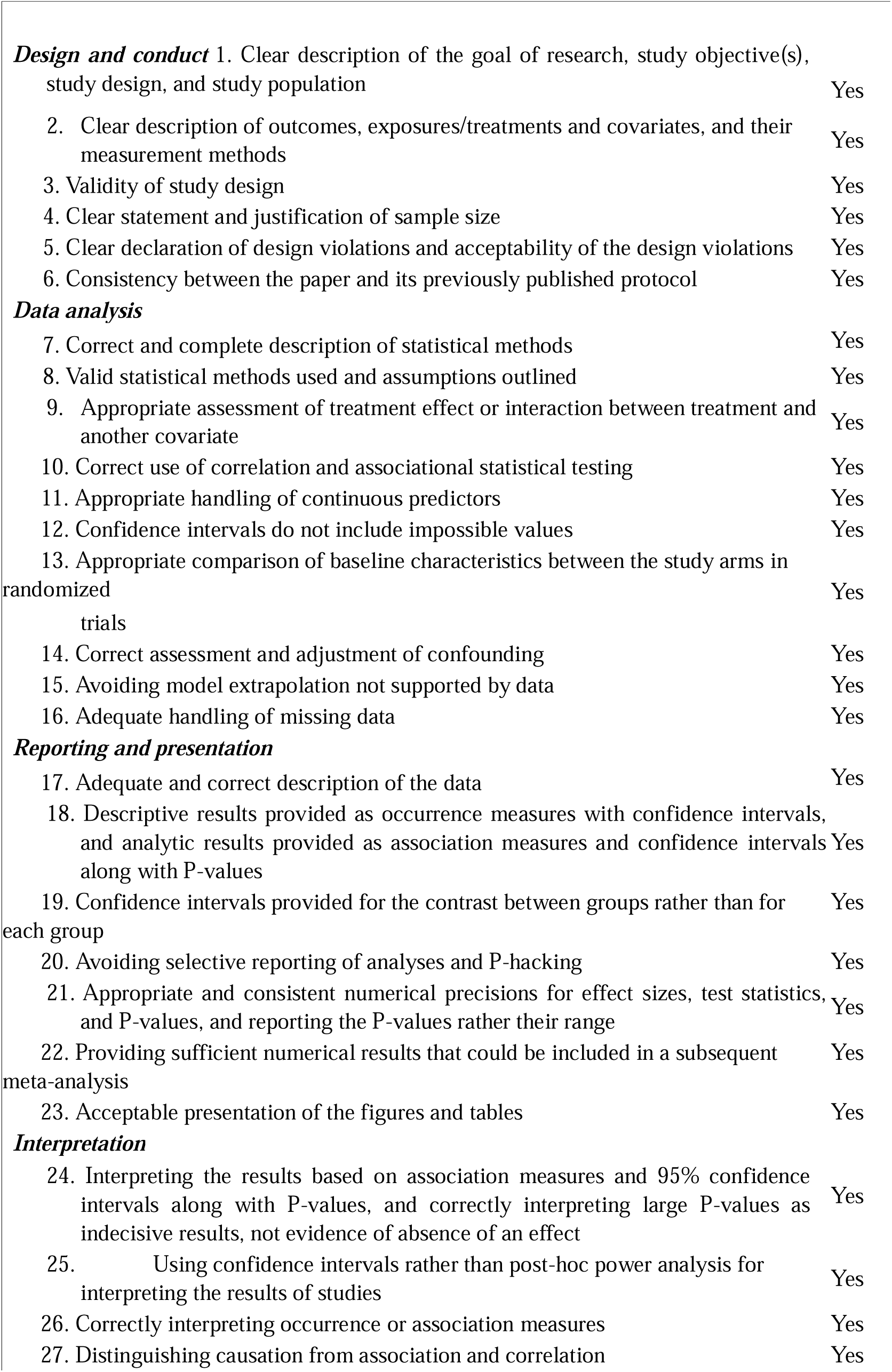
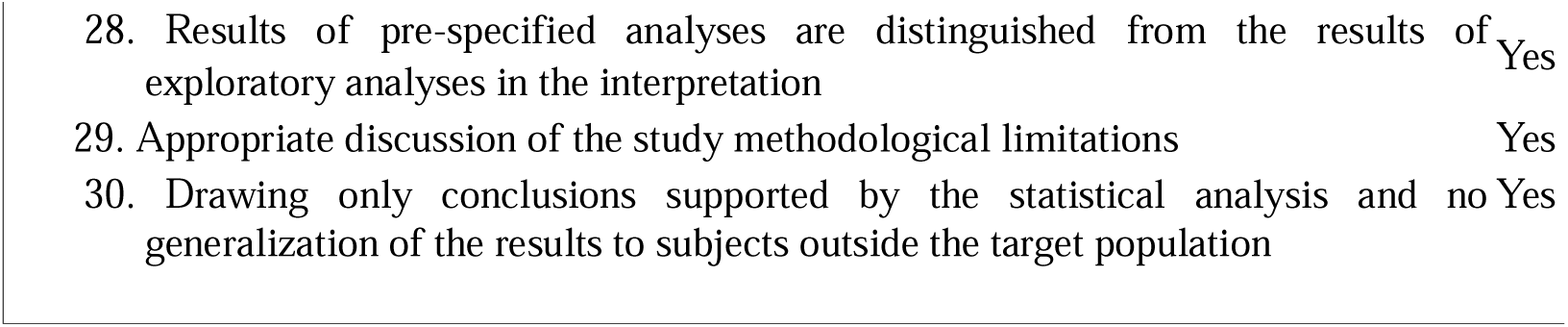

